# Improved prediction of blood biomarkers using deep learning

**DOI:** 10.1101/2022.10.27.22281549

**Authors:** Arnór I. Sigurdsson, Kirstine Ravn, Ole Winther, Ole Lund, Søren Brunak, Bjarni J. Vilhjálmsson, Simon Rasmussen

## Abstract

Blood and urine biomarkers are an essential part of modern medicine, not only for diagnosis, but also for their direct influence on disease. Many biomarkers have a genetic component, and they have been studied extensively with genome-wide association studies (GWAS) and methods that compute polygenic scores (PGSs). However, these methods generally assume both an additive allelic model and an additive genetic architecture for the target outcome, and thereby risk not capturing non-linear allelic effects nor epistatic interactions. Here, we trained and evaluated deep-learning (DL) models for PGS prediction of 34 blood and urine biomarkers in the UK Biobank cohort, and compared them to linear methods. For lipid traits, the DL models greatly outperformed the linear methods, which we found to be consistent across diverse populations. Furthermore, the DL models captured non-linear effects in covariates, non-additive genotype (allelic) effects, and epistatic interactions between SNPs. Finally, when using only genome-wide significant SNPs from GWAS, the DL models performed equally well or better for all 34 traits tested. Our findings suggest that DL can serve as a valuable addition to existing methods for genotype-phenotype modelling in the era of increasing data availability.

## INTRODUCTION

Biomarkers found in blood and urine play an important role in the diagnosis and monitoring of disease, such as cardiovascular, kidney and liver disease^1–3^. For example, evidence points to lipoprotein(a) being a causal factor for cardiovascular disease^4^, and it has been associated with increased risk of thrombosis and stroke^5,6^. Furthermore, they have been used to assess treatment response^7^, prognosis^8^, as a measure of drug efficiency^9^, drug development^10^ and for disease/outcome prediction^11,12^. Many biomarkers have a genetic component, which has been shown with genome-wide-association-studies (GWAS)^13–15^, and methods that compute polygenic scores (PGSs)^16^. Therefore, a method that better predicts biomarkers levels such as lipoprotein(a) from genotype data can be expected to better stratify patients regarding cardiovascular disease risk. In addition to better patient stratification, analyzing the factors that drive the increased performance of non-linear models could uncover valuable biological insights. For example, improved understanding of the genetic mechanism of traits that contribute to disease might assist in the development of new drugs and therapies.

PGSs combine the effect of many genetic variants on a phenotype, which can either be qualitative (e.g., disease status) or quantitative (e.g., blood biomarker level). While ethical and societal implications need careful consideration before widespread deployment^17,18^, PGSs are increasingly being considered for their clinical utility, e.g., in the context of precision medicine^17,19–23^. Numerous methods exist for computing trait and disease PGSs on individual-level data^24–28^ and summary statistics^29–34^, but they generally only model additive relationships between genotype and target. For blood biomarkers in particular, recent studies have successfully shown they can be partially predicted with additive methods^16^. Non-linear models have had mixed success in human genomic prediction^35,36^, but recent methods are starting to show promise^37–39^. We hypothesized that using deep learning (DL) based PGS predictions, it would be possible to identify non-linear effects on blood and urine biomarkers. Furthermore, we wanted to investigate whether an improved *R*^2^ was due to improved modeling of covariates, proper modeling of recessive and dominant variants, and genetics interactions.

Here, we trained additive and deep learning models to compute PGSs for the levels of 34 blood and urine biomarkers, using individual-level data in the UK Biobank^40^. When using around 500K SNPs, we found that on lipid traits, the DL models greatly outperformed state-of-the-art additive models, which we found consistent across different populations. We analyzed the factors driving the DL performance gain, and found them to be due to non-linear effects in age and sex, non-additive effects of genotypes, and interaction effects between different SNPs (e.g., possibly due to epistasis). Furthermore, when restricting the analysis to 9K SNPs identified with GWAS, the DL models performed equally well or better on all 34 traits. This work demonstrates that DL is a viable option for PGSs, as it can improve prediction substantially over existing methods. We expect DL to become increasingly common within genetics as data availability grows, where it might not only better stratify patients when it comes to risk, but also lead to new biological discoveries and applications.

## RESULTS

### Deep Learning for Polygenic Prediction of Biomarkers

In this work, we examined the contribution of complex non-linear effects in the polygenic prediction of 34 biomarkers measured in blood and urine. Compared to diseases, one benefit of predicting biomarkers is that they have been measured for many samples, which increases the likelihood of the models identifying complex effects assuming such effects are present (Supplementary Data 1). Furthermore, instrumentally measured biomarker levels may provide a better signal when training the PGS models compared to, for example, a broad disease diagnosis. Using 488,377 individuals and 803,113 SNPs from the UK Biobank cohort^40^, we performed basic quality-control (QC) steps (see Methods) and then split our data into training/validation and test sets. Then, we compared the DL model performance to two state-of-the-art methods for PGSs, `bigstatsr`^26^ and `snpnet-2.0`^28^. For those traits where the DL model performed better, we examined whether the gap was driven by the covariates age and sex or if it was due to complex genotype effects. Here, we found the effects coming from both sources, and within the genotype data we found both non-additive and interaction effects (Figure 1).

**Figure 1.**
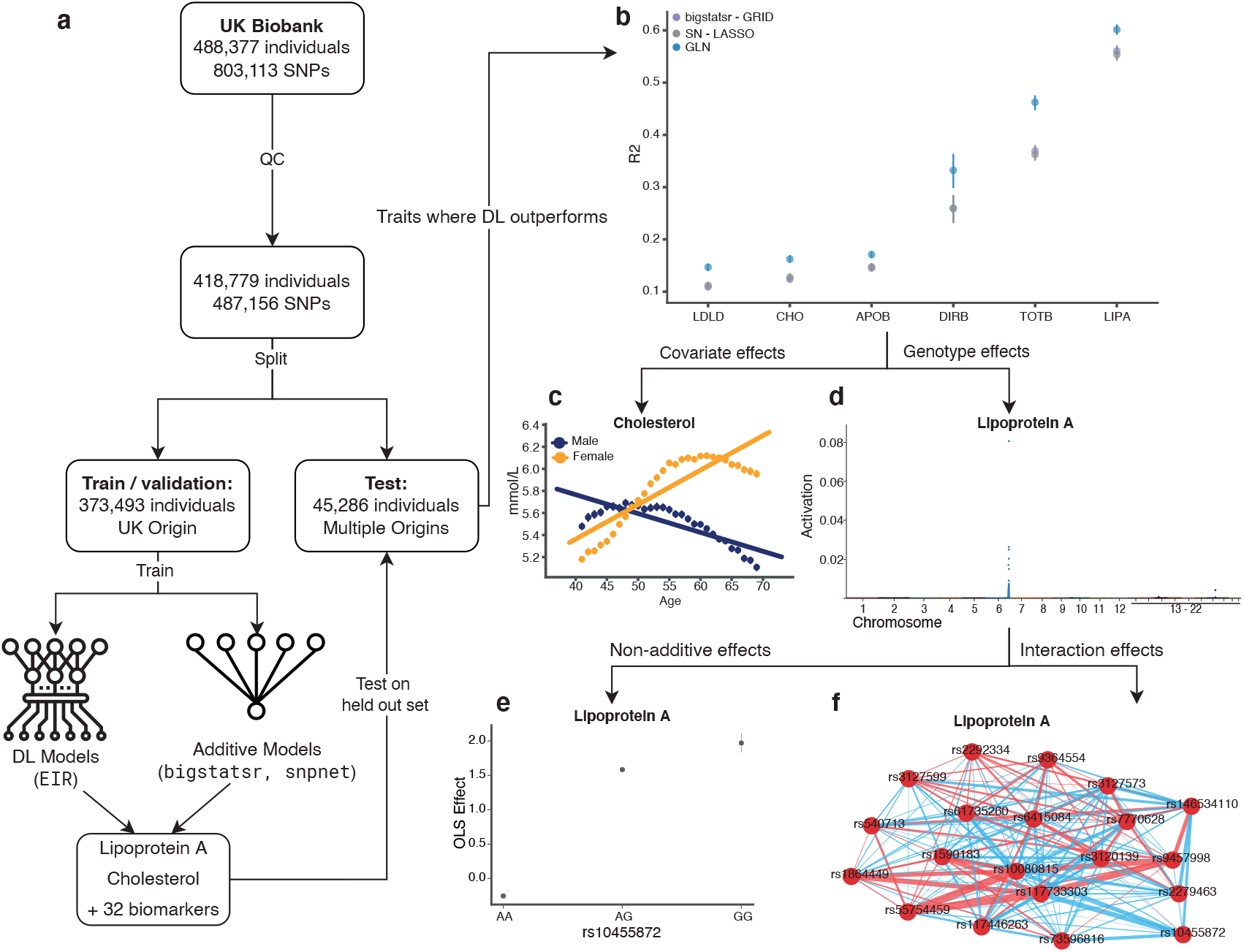
Study Overview. **(a)** A flow diagram showing the high-level steps taken for the model training and evaluation. **(b)** After training, the deep-learning based genome-local-net (GLN) model (blue) performed particularly well for six lipid traits compared to the additive `bigstatsr` (purple) and `snpnet-2.0` (gray) models. Performance shown on samples with self-reported UK origin. Bars represent the 95% CI from 1,000 bootstrap replicates on the held-out test set. **LDLD**: LDL Direct, **CHO**: Cholesterol, **APOB**: Apolipoprotein B, **DIRB**: Direct Bilirubin, **TOTB**: Total Bilirubin, **LIPA**: Lipoprotein(a). **(c-f)** Examining the traits where GLN outperformed the linear models, we found non-linear effects in the covariates (c) and genotype data (d). For the genotype data, we found both non-additive effects (e) and interaction effects (f).

### Deep learning outperforms additive models for biomarker prediction

We trained and evaluated all models on two sets of genotype data. The first set (denoted “full set”) used 487,156 SNPs resulting from the QC. When training on this set, we found the DL-based model to overfit on some traits based on validation set performance. Therefore, we also used a combined set of 9,034 SNPs that had a GWAS hit with any of the biomarkers when performed on the training/validation set (denoted “GWAS set”) (Supplementary Data 2). Such a pre-filtering approach, using both linear and non-linear approaches, has been successfully applied in previous work^37,38^. Across the 34 biomarkers, we found that the DL-based genome-local-net^37^ (GLN) model performed particularly well for cholesterol, LDL cholesterol, apolipoprotein B, direct bilirubin, total bilirubin and lipoprotein(a) compared to the additive models in both sets (Figure 2a,b). For the full set, we found the differences to range between 0.024 (apolipoprotein B) to 0.093 (total bilirubin) *R*^2^ gain when compared to the best performing additive models (Supplementary Data 3). For the GWAS set, the difference increased even further to 0.034-0.102 *R*^2^ (Supplementary Data 4). In the full set, we also found examples of the DL models performing worse than the best performing additive models. The biggest gaps were for HDL cholesterol (−0.029 *R*^2^), glycated haemoglobin HbA1c (−0.037 *R*^2^) and gamma glutamyltransferase (−0.047 *R*^2^). The decrease in performance was most likely explained by the DL models overfitting on the full set, indicated by divergence in training and validation set performances. However, as noted above, for the GWAS set, the DL models performed equally well or better than the additive models for all traits. For example, the trait where the additive models had the highest increase compared to the DL models, HDL cholesterol, only had a difference of -0.008 *R*^2^. This strongly indicates that DL models can capture all the variance additive models can, and more, on biobank scale genotype data. However, when modelling on such a large input space, we might not be in a high enough data availability yet for the current DL model implementations to consistently outperform. For some traits, the chosen GWAS inclusion threshold of (5 *×* 10^−8^)*/*34 was likely too strict, as the GWAS set had worse performance, for both GLN and linear models, as compared to the full set (Supplementary Figure 2). Finally, we extended our analysis to more diverse groups of people than those originating from the UK. Here, we found the increased performance of the DL models to be consistent across non-UK European, African, and Asian continents of origin. This was the case for all six lipid traits, and across both the full set and GWAS set of SNPs. The gains ranged from 0.0037 (cholesterol, African continent of origin, GWAS set) to 0.12 *R*^2^ (lipoprotein(a), Asian continent of origin, GWAS set) (Figure 2c-e, Supplementary Figure 3 and Supplementary Data 3).

**Figure 2.**
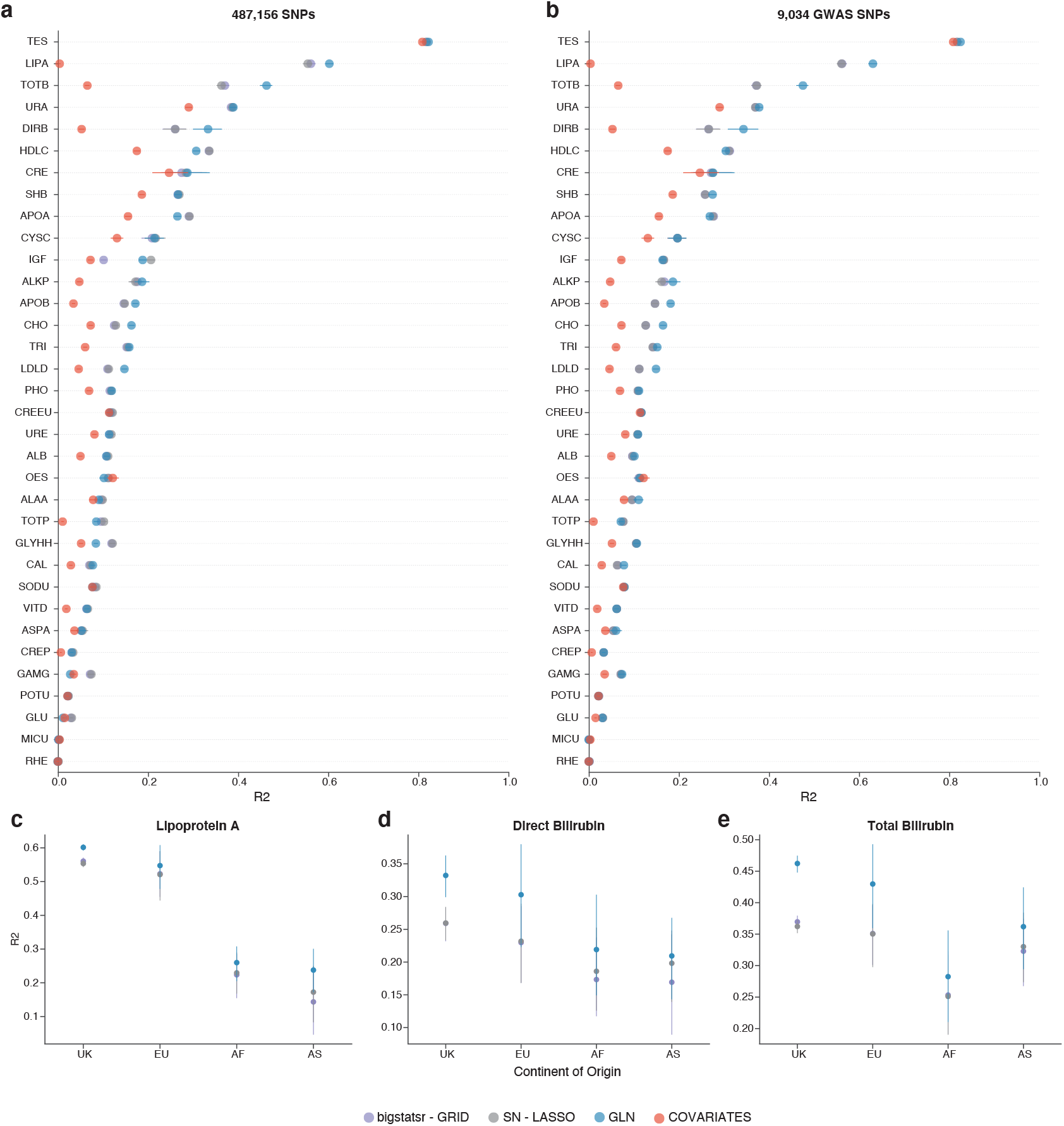
Model Performance. Comparison of deep-learning (DL) based genome-local-net (GLN) model (blue), additive `bigstatsr` model (light purple), additive `snpnet-2.0` (SN) LASSO model (gray) and covariate based neural-network models (red) performance on samples of UK origin within the held-out test set, measured in *R*^2^. All models were adjusted for age, sex, and the first 10 genomic principal components. **(a)** Models trained on a set of 487,156 SNPs after general QC. **(b)** Models trained on a set of 9,034 SNPs identified by performing a genome-wide-association-study (GWAS) on the training and validation sets for all target biomarkers. **TES**: Testosterone, **LIPA**: Lipoprotein(a), **TOTB**: Total Bilirubin, **URA**: Urate, **DIRB**: Direct Bilirubin, **HDLC**: HDL Cholesterol, **CRE**: Creatinine, **SHB**: Sex hormone-binding globulin, **APOA**: Apolipoprotein A, **CYSC**: Cystatin C, **IGF**: IGF1, **ALKP**: Alkaline Phosphatase, **APOB**: Apolipoprotein B, **CHO**: Cholesterol, **TRI**: Triglycerides, **LDLD**: LDL Direct, **PHO**: Phosphate, **CREEU**: Creatinine Enzymatic in Urine, **URE**: Urea, **ALB**: Albumin, **OES**: Oestradiol, **ALAA**: Alanine Aminotransferase, **TOTP**: Total Protein, **GLYHH**: Glycated Haemoglobin HbA1c, **CAL**: Calcium **SODU**: Sodium in Urine, **VITD**: Vitamin D, **ASPA**: Aspartate Aminotransferase, **CREP**: Creactive Protein, **GAMG**: Gamma Glutamyltransferase, **POTU**: Potassium in Urine, **GLU**: Glucose, **MICU**: Microalbumin in Urine, **RHE**: Rheumatoid Factor. **(c-d)** Performance in *R*^2^, stratified by self-reported continent of origin on the 487,156 SNP set. **UK**: United Kingdom, **EU**: Non-UK Europe, **AF**: Africa, **AS**: Asia. Bars represent the 95% CI from 1,000 bootstrap replicates on the held-out test set.

### Non-linear patterns in covariates better explain biomarker levels

Knowing that the DL models were performing favorably, we decided to examine what was driving the performance gain over the additive models, focusing on the six lipid traits. First, we investigated the biomarker levels as a function of age and sex, and found lipoprotein(a), total bilirubin and direct bilirubin to be either poorly explained or have a mostly linear pattern (Figure 3d-f). However, we found apolipoprotein B, LDL direct and cholesterol to all share a similar, non-linear pattern (Figure 3d-f). To examine more concretely how these patterns were reflected in model performance, we trained and compared a linear model to a non-linear DL model, both using only the covariates as inputs. Here, we found the performance gain of the non-linear model closely followed linear and non-linear patterns. This indicates that for apolipoprotein B, LDL direct and cholesterol, non-linear effects in the covariates strongly contribute to the improved performance of DL compared to additive genotype models (Figure 3). Extending the analysis to the other 28 biomarkers, we identified non-linear effects in 19 out of the 34 biomarkers (Supplementary Figure 1). While it is trivial to model the effect by adding the correct terms to a linear model (e.g., age^2^), one advantage of NN based models is their ability to find and utilize such effects automatically. This can be advantageous when modelling on high dimensional data more abstract than age, such as protein expression, where widespread complex effects might be present.

**Figure 3.**
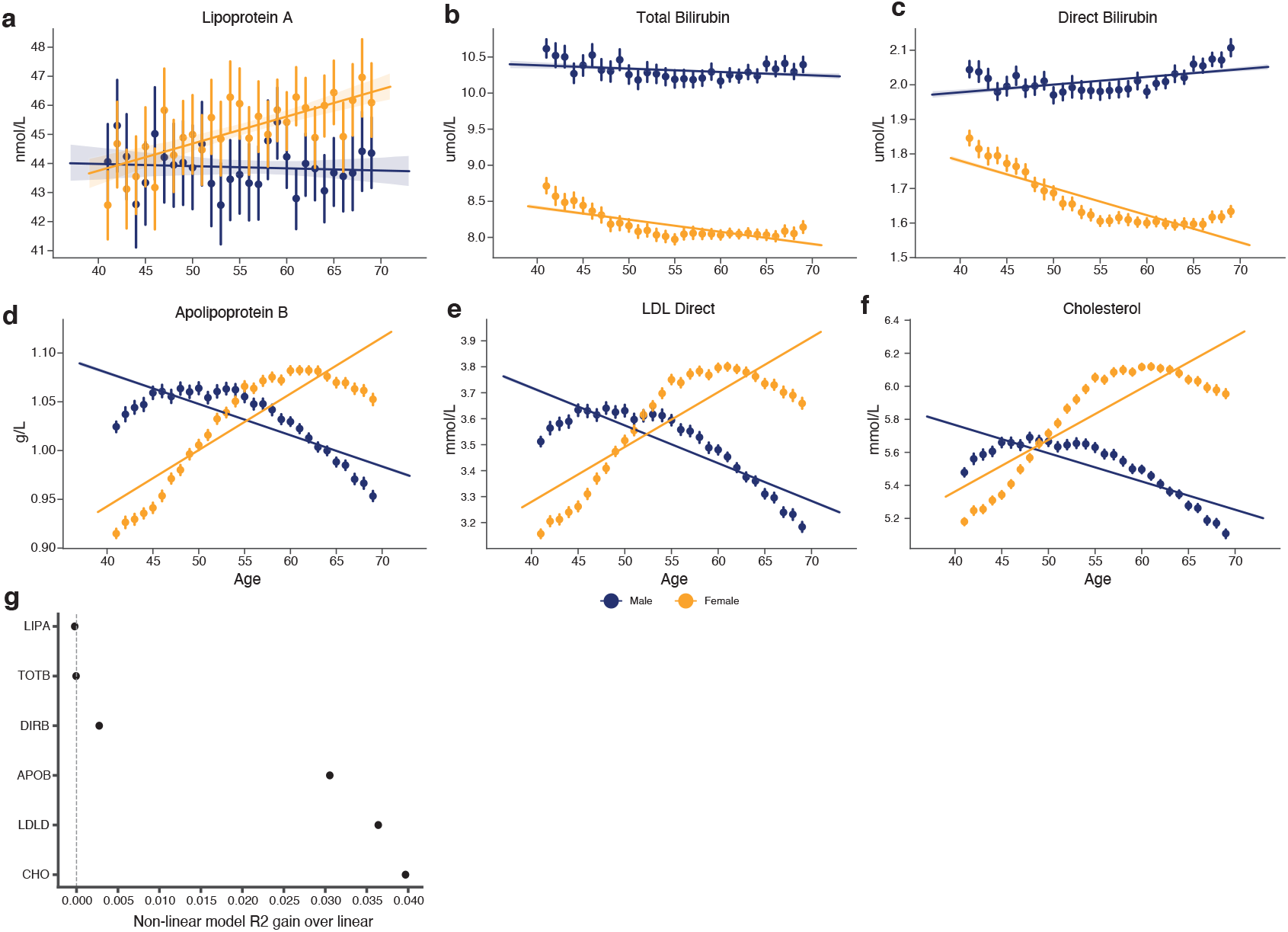
Effect of age and sex on biomarker levels. **(a-f)** Six traits where the DL models outperformed the additive models, showing both linear and non-linear patterns. The levels are plotted as a function of age, stratified by males (dark blue) and females (orange). Bars represent the 95% confidence interval after 1,000 bootstrap iterations. **(g)** *R*^2^ performance gain of training a non-linear DL models over linear models using covariates as inputs. **LIPA**: Lipoprotein(a), **TOTB**: Total Bilirubin, **DIRB**: Direct Bilirubin, **APOB**: Apolipoprotein B, **LDLD**: LDL Direct, **CHO**: Cholesterol.

### Biomarkers levels are influenced by non-additivity in the genome

Next, we examined the contribution of non-additive effects from alleles, such as dominance. First, we, for each trait, used SHAP^41,42^ to identify the top 100 most activated SNPs by the DL GLN model on the validation set (Supplementary Figure 4 and Supplementary Data 5). To investigate the effects, we fitted an ordinary least squares (OLS) model on the SNPs in question and found multiple significant (*p <* 0.05*/*(100 *×* 34)) examples of non-additive effects (Supplementary Table 1 and Supplementary Data 6). Two interesting cases were rs10455872 (chr6, LPA gene), which showed a dominance effect for lipoprotein(a) and rs887829 (chr2, UGT1A1 gene), which had a recessive effect on total bilirubin. Next, we examined the effect of these SNPs on the additive and DL model output distribution. The effects on the output indicate how, and to what extent, the models are using particular genotypes to generate a prediction. As expected, we found the output distribution of the additive `bigstatsr` model to be strongly modulated by the genotypes of these SNPs in an additive pattern. However, for the DL models, we found that the output distributions were much more closely linked to the effects identified by the OLS model (Figure 4). When investigating the literature, we found that rs10455872 is a well-known SNP that has been associated with lipoprotein(a) levels and coronary disease^43,44^. Additionally, rs887829 has been strongly linked to variation in bilirubin levels and Gilbert’s syndrome^45^, a genetic condition known for autosomal recessive inheritance. These results indicate that measured levels of many biomarkers are strongly modulated by non-additive effects, and compared to models which impose an additive prior on alleles, one type of effect DL models utilize to better predict their levels.

**Figure 4.**
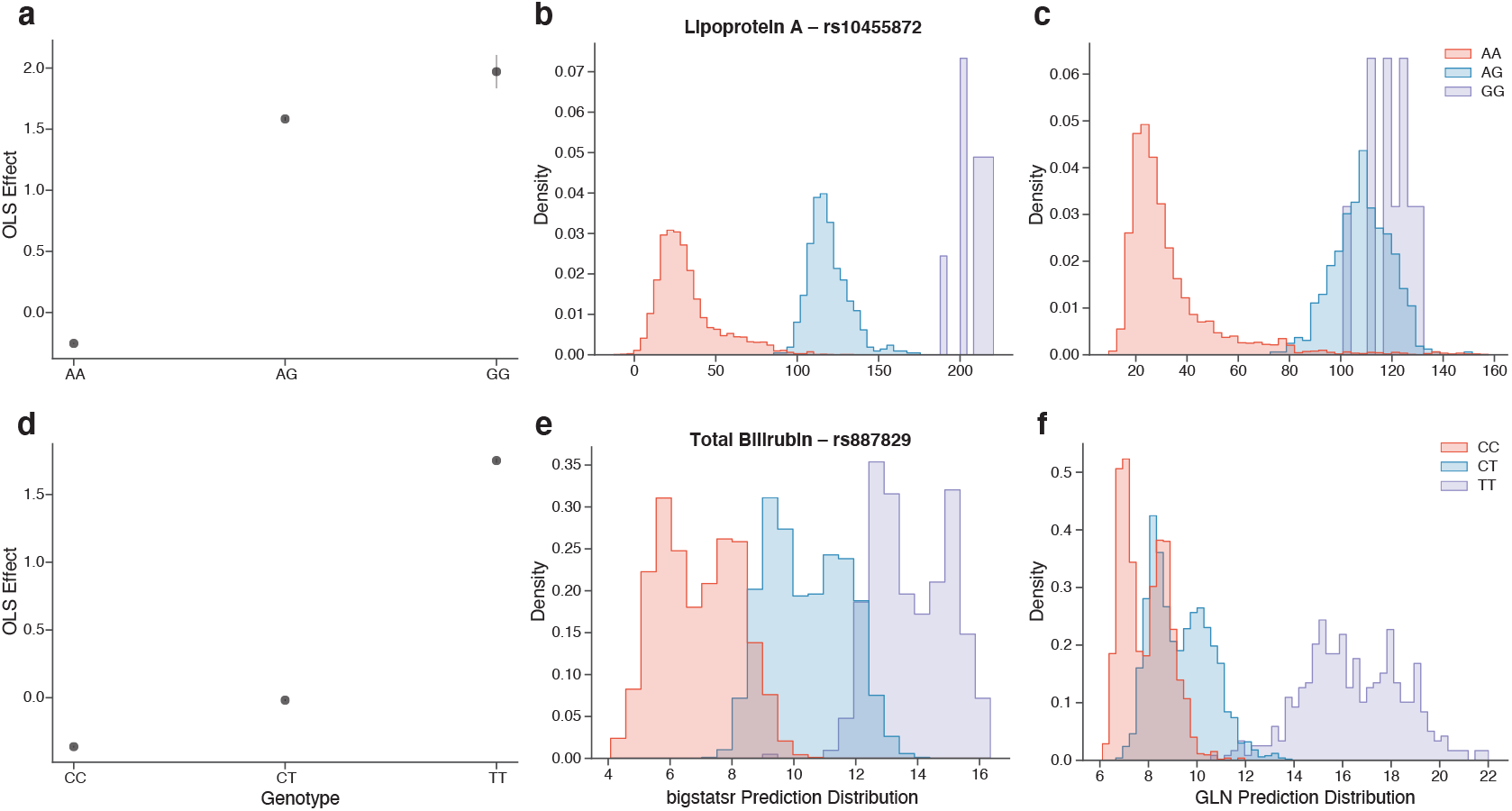
Deep learning detects and utilizes non-additive effects. **(a, d)** Effects from fitting an ordinary least squares (OLS) model showing non-additive dominant (top row) and recessive (bottom row) effects of SNPs modulating the effects of lipoprotein(a) and total bilirubin. Bars represent the 95% confidence interval of the estimated parameters for each genotype. **(b, e)** Output distribution of additive `bigstatsr` model colored by genotype (red: homozygous reference, blue: heterozygous, light purple: homozygous alternative). **(c, f)** Output distribution of non-linear genome-local-net (GLN) model colored by genotype (red: homozygous reference, blue: heterozygous, light purple: homozygous alternative). The output distribution of the GLN models indicates that the model captures both dominant and recessive effects.

### Level of lipoprotein(a) is influenced by interaction effects

Knowing that non-linear covariate effects and non-additive effects were both contributing to the better performance of the DL models, we next examined the contribution of interaction effects in the genotype data (e.g., due to epistasis). First, we examined the effect of allowing linear models to model on genotypes separately, allowing them to capture effects such as dominance. Here, we trained three types of linear models and one non-linear model on genotype data encoded in an additive (i.e., an SNP takes values 0-2 for homozygous reference, heterozygous or homozygous alternative genotypes, respectively) and one-hot (i.e., homozygous reference, heterozygous or homozygous alternative genotypes are each assigned a parameter/weight separately) format. We used Ridge, Lasso, and Elastic Net regression as the linear models, while gradient boosted decision trees (GBTREE) were used as a non-linear model. As general implementations of these models are not expected to scale to high dimensional genotype data, we used the subset of SNPs (Supplementary Data 7) that had the highest activations by the DL GLN model (defined as average absolute SHAP values on the validation set). Using both additive and one-hot data encoding allows us to examine if the difference between additive and DL models can be closed by simply removing the additive assumption w.r.t. alleles. For direct and total bilirubin, we found that the gap was mostly closed by using linear models with one-hot encoded genotype data, indicating that non-additive effects such as dominance were the largest driver (Figure 5a,b and Supplementary Data 8). However, lipoprotein(a) showed only a small increase in performance when using one-hot encoded data, but a large gap when using the non-linear model, indicating possible complex effects from the genotype data (Figure 5c, Supplementary Data 8). Additionally, these patterns were consistent across UK, Non-UK European, African, and Asian continents of origin (Supplementary Figure 5-8, Supplementary Data 8). To confirm and examine the effects further, we computed all pairwise interactions for the top 100 DL activated SNPs for each trait, and found multiple (*p <* 0.05*/*(100^2^ *×* 34)) such effects, particularly for the six lipid traits where DL outperformed (Figure 5d-f, Supplementary Figure 9, Supplementary Data 9). For instance, lipoprotein(a), total bilirubin and direct bilirubin had particularly numerous and strong interaction effects compared to the other traits (Supplementary Data 9). Among these, lipoprotein(a) showed the strongest effects, and has previously been associated with epistatic effects^46^ (Figure 5e, Supplementary Figure 9, Supplementary Data 9).

**Figure 5.**
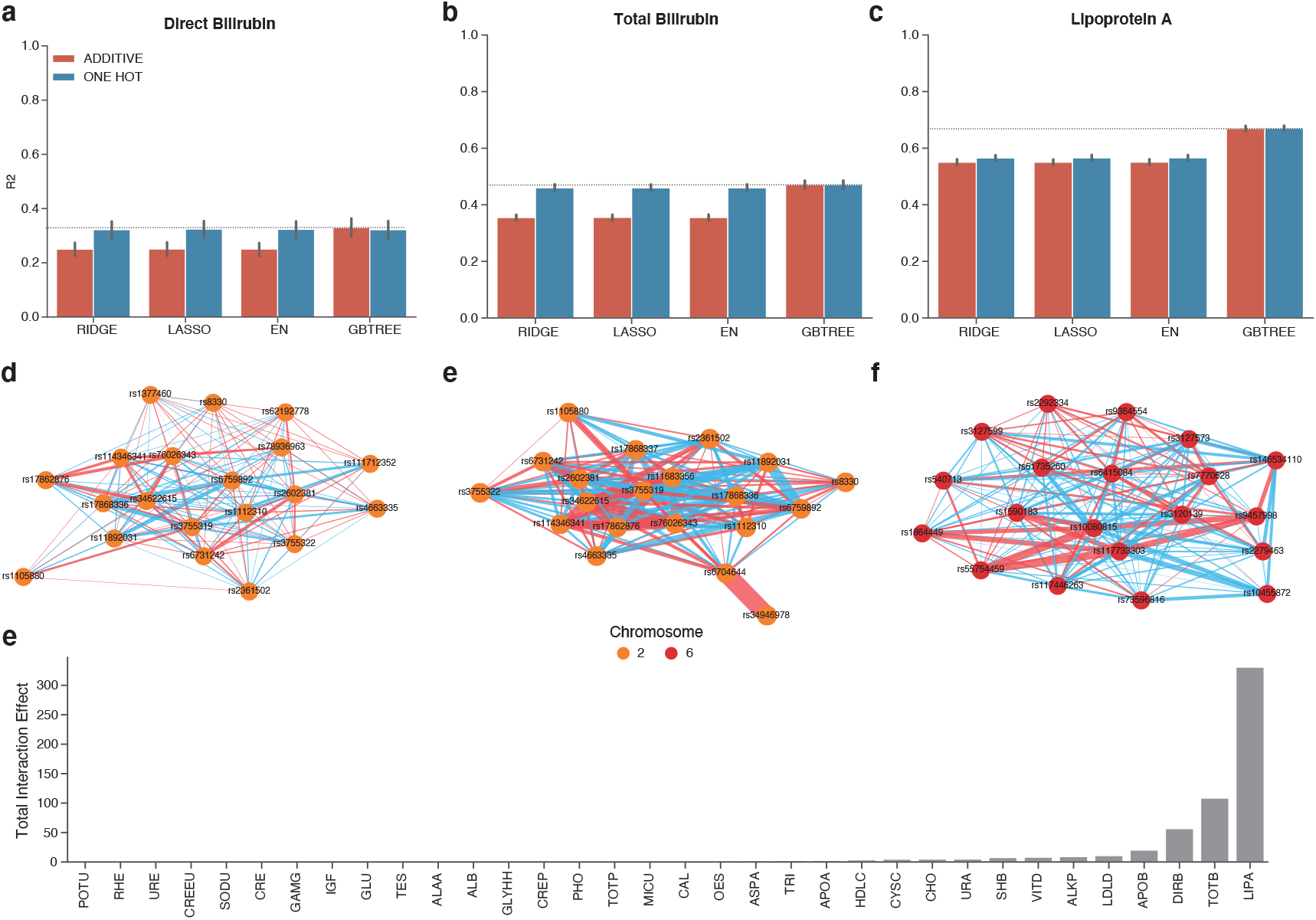
Interaction effects. **(a-c)** Performance comparison of linear and non-linear models, trained on samples of self-reported UK origin, evaluated on samples with self-reported UK origin. For each trait, a subset of SNPs most highly activated by the genome-local-net (GLN) model on the validation set were used for training and evaluation. All models were trained on data encoded with an additive prior (dark blue) and in a one-hot encoding (red), where each genotype can be modelled separately. Bars represent the 95% CI from 1,000 bootstrap replicates on the held-out test set. **RIDGE**: Ridge Regression, **LASSO**: Lasso Regression, **EN**: Elastic-Net regression, **GBTREE**: Gradient Boosted Decision Trees Regression. **(d-e)** Computing the interactions between SNPs with an ordinary-least-squares (OLS) model finds numerous interaction effects, which vary in strength and number depending on trait. Shown are the top 20 SNPs with the highest total interaction effects for each trait. Each node represents an SNP, and the edges reflect interaction effects identified by the OLS model affecting a biomarker level. The edges a colored according to whether the interaction effect increases (red) or decreases (blue) the biomarker level, and the width of the edges reflect the relative strength of the interaction between SNPs. Nodes are colored by chromosome (orange: 1, red: 6, teal: 19). **(e)** Total SNP interaction effect for each trait found by summing up individual effects identified by OLS. **POTU**: Potassium in Urine, **RHE**: Rheumatoid, **URE**: Urea, **CREEU**: Creatinine Enzymatic in Urine, **SODU**: Sodium in Urine, **CRE**: Creatinine, **GAMG**: Gamma Glutamyltransferase, **IGF**: IGF1, **GLU**: Glucose, **TES**: Testosterone, **ALAA**: Alanine Aminotransferase, **ALB**: Albumin, **GLYHH**: Glycated Haemoglobin Hba1c, **CREP**: Creactive Protein, **PHO**: Phosphate, **TOTP**: Total Protein, **MICU**: Microalbumin in Urine, **CAL**: Calcium, **OES**: Oestradiol, **ASPA**: Aspartate Aminotransferase, **TRI**: Triglycerides, **APOA**: Apolipoprotein A, **HDLC**: HDL Cholesterol, **CYSC**: Cystatin C, **CHO**: Cholesterol, **URA**: Urate, **SHB**: SHBG, **VITD**: Vitamin D, **ALKP**: Alkaline Phosphatase, **LDLD**: LDL Direct, **APOB**: Apolipoprotein B, **DIRB**: Direct Bilirubin, **TOTB**: Total Bilirubin, **LIPA**: Lipoprotein A.

## DISCUSSION

Here, we show that DL can greatly improve predictive performance for blood biomarkers, particularly lipid traits, compared to the state-of-the-art additive models. For the 6 lipid traits where DL had a clear advantage, three traits had non-linear covariate effects, two strong non-additive effects and one strong interaction effects between SNPs. Even though performance of the all models were lower when applied to non-UK populations, the improvement of using the DL models were still consistent. Therefore, the underlying factors driving DL model advantages for these biomarkers are likely conserved across ethnicities. This indicates that the DL models are likely learning biological effects and not cultural differences. Future work includes a more thorough examination of the identified effects, as well as examining whether gene-environment effects contribute to the DL performance gain.

The improvement of using DL models was even more evident when reducing the number of SNPs included in the model, where the DL models performed equally well or better for all the 34 traits tested. This indicates that complex effects are present in more than the six lipid traits. For example, in the GWAS set, the DL models additionally outperformed all linear models on testosterone, urate, sex hormone-binding globulin, alkaline phosphatase, triglycerides, alanine aminotransferase and calcium. One drawback of using GWAS results, or those from any linear model, to pre-select SNPs is that it will likely miss SNPs that exclusively have non-linear effects. Alternatively, one could use the SNPs identified by a DL model on the training/validation set, which might enrich the set of used SNPs. Another factor is that choosing which SNPs to include introduces another complexity in the experiment design, i.e., which criteria and thresholds to use for inclusion. Methods based on linear models generally approach this by iteratively testing multiple penalization values, and choosing a value that performs best on the validation set. Such an approach can just as well be applied to DL-based models, which might provide a future area of research.

However, with more data becoming available, one might expect less tuning, particularly regarding regularization, to be needed. This was particularly evident on the GWAS set, where the DL models performed equally well or better than the carefully tuned linear models, for all traits. The fact that such a gain was seen on the GWAS set indicates that for high dimensional genotype data, we might be rapidly approaching a level of data availability where DL consistently outperforms other methods in predictive performance, which has already happened in the fields of computer vision and natural language. Performance is not all, however, as interpretable models will be essential for clinical applicability of PGSs. Therefore, DL can be viewed as an additional tool in the algorithmic toolbox, identifying traits and inputs where complex effects play a role. Ongoing and future work therefore includes examining whether the identified genotype effects, additive or not, can be better connected with the biology of the traits in question^47^.

As the PGSs become increasingly relevant for the clinic, methods that better predict factors, which can have causal effects on disease, provide a clear value for patient stratification and disease prevention. An additional benefit of the DL methods are their inherent capabilities to integrate multi-modal data. In the future, personalized medicine will likely be realized by combining genomics, proteomics, imaging, textual descriptions, biochemical measurements and various other factors that describe or contribute to a person’s medical state and trajectory^37,48^. Therefore, methods that allow for such multimodal integration are expected to become pervasive in personalized medicine.

## METHODS

### Processing of UK Biobank genotype and clinical data

The genotype data was processed using Plink^49^, version v1.90b6.10. After processing, the genotype data was converted to 452,976 sample arrays with 487,156 SNPs each, encoded in one-hot format (i.e., 4 values per SNP). We used the following parameters in Plink, `--maf 0.0001 --geno 0.03 –mind 0.1 --hwe 1e-10` as well as removing samples with a kinship of more than 0.1 and finally performing LD pruning with `--indep-pairwise 1500 150 0.8` in Plink. Only autosomal chromosomes were included. For the GWAS, we used the implementation from the `bigsnpr` tool^26^, with a p-value threshold of (5 *×* 10^−8^)/34. Unless otherwise specified, age, sex and the first 10 genotype principal components were included during training. For the tabular data, continuous columns were standardized using the training set statistics in all experiments, meaning that the values computed for the training set were applied to the validation and test sets. Missing values were imputed with the averages from the training set. Categorical columns were numerically encoded, missing values were marked as “NA” before numerical encoding. Performance on the held-out tests set are reported as the average and 95% CIs after performing 1,000 bootstrap replicates, following a similar approach as applied before^33^.

### Training implementation and approach

All NN models were implemented using Pytorch^50^, version 1.10. A held-out test set was used for all models to get a final performance after training and evaluating on train and validation sets, respectively. On the training/validation sets, we did a 3-fold Monte Carlo cross-validation (CV) with a final ensemble prediction on the test set. The continuous biomarker target values were normalized with statistics computed on the training set. We used mean squared error loss during training for the regression tasks. All models were trained with a batch size of 64. During training, we used plateau learning rate scheduling to reduce the learning rate by a factor of 0.2 if the validation performance had not improved for 10 steps, with a validation interval of 500 steps. We used early stopping to terminate training when performance had not improved for a certain number of validation steps, with patience of 16 steps in all cases. The early stopping criterion was activated after a buffer of 2,000 iterations. All models were trained with the Adam optimizer^51^ with a weight decay of 1 *×* 10^−4^ and a base learning rate of 1 *×* 10^−4^. All neural network architectures used the SiLU^52,53^ (also known as Swish^54^) activation function with a trainable parameter *β* inside the sigmoid function. When using weight decay, we did not apply it to the *β* parameter. We also used a gradient clipping of 1.0 during training. For the neural network models, we augmented the genotype input by randomly setting 20% of the SNPs as missing in the one-hot encoded array. We further augmented the genotype data by applying a block CutMix^55,56^ augmentation with a mixing *α* settings of [0.1, 0.2, 0.4] for the CV folds 1, 2 and 3 respectively. All NN models were trained on a single 16 GB NVIDIA® V100 Tensor Core GPU. The exact `EIR` configurations (for version `0.1.24a0`) used for model setup and training are made available (Supplementary Data 10).

### Comparison to benchmark methods

The `bigstatsr` training was done using a 3-fold CV using a grid search *α* = [0.0001, 0.001, 0.01, 0.1, 1] for the elastic net mixing parameter, and the method additionally tests various values for the *λ* penalization parameter. The method then performs an ensemble-like procedure across the folds to produce the final model, which is evaluated on the test set. For `snpnet-2.0`, Ridge, Lasso, and Elastic Net models were fit on the training set and used the validation set to find the optimal *λ* penalization parameter. The models were trained with 2000 SNPs per batch, 100 iterations, 20 *λ* values in the first iteration, 10 extended *λ* values and a convergence threshold of 1 *×* 10^−7^. All `snpnet-2.0` models were then re-fit on the training and validation set together using the identified optimal *λ*. We only report the best performing `snpnet-2.0` model, the Lasso variant, in the main results, but the results for Elastic Net and Ridge variants are available in the supplementary data (Supplementary Data 2).

### Investigation of non-linearity

To analyze the effect of data encoding and non-linearity on predictive performance, we used the Ridge, Lasso, and Elastic Net implementations from `scikit-learn` and gradient boosted decision trees implementation from `XGBoost`^57^. To select which SNPs were included in the modelling for each trait, we used a goodness-of-fit test to identify a distribution best fit of the DL GLN model SNP activations on the validation set. After identifying the distribution, we used a p-value threshold of 0.05*/*487156 (denominator refers to total number of SNPs). After p-value thresholding, additional minimum and maximum cutoffs of 128 and 512 were applied for the number of SNPs to include, per trait. Additionally, we included the same covariates as used to train the previous models. For the linear models, we performed a 5-fold CV on the training/validation set. For the Lasso model, we tested 100 values for *λ* with *ε* = 1 *×* 10^−7^. The Elastic Net use the same parameters, but we also tried values [0.1, 0.5, 0.7, 0.9, 0.95, 0.99] for the mixing parameter. The Ridge regression used the default parameters as defined by `scikit-learn`. The gradient boosted decision trees used a learning rate of 0.02, maximum depth of 7, 10,000 boosting iterations, early stopping based on the validation set and a 100% training set subsample for each boosting iteration. The same training, validation and tests sets were used as for the GLN model training and evaluation.

### Deep Learning Architectures

All NN models use the genome-local-net (GLN) architecture^37^ for the genotype feature extraction, which uses residual blocks^58^ with locally connected layers (LCLs)^59–61^ as a backbone. In the first LCL, we used a kernel width of 4 (covering one SNP per group) and 4 output sets. In the subsequent four residual blocks, we used a larger kernel width of 16 and 4 output sets for the first two blocks and a kernel width of 16 and 8 output sets in the last two blocks. We used a small L1 penalization of *λ* = 1 *×* 10^−6^ in the first LCL layer. We used a dropout^62^ of 0.25 in the LCL feature extractor residual blocks. The tabular feature extractor used in all models used embeddings for categorical inputs and normalized continuous values with statistics from the training set. The tabular inputs were then concatenated and passed through a single FC layer. The fusion module aggregated the intermediate representations from the individual feature extractors by concatenating. The final feature sizes for the genotype and tabular feature extractors were 1904 and 14 respectively, which resulted in a total feature size of 1918 passed to fully connected residual layers. The FC layer dimension in the FC blocks was set to 1,024. Additionally, we increased the dropout to 0.5 in those layers and added stochastic depth^63^ with a survival rate of 0.8. After the final residual blocks, there was a set of `BN-ACT-DO-FC` layers which computed the final output for a given trait. The exact `EIR` configurations (for version `0.1.24a0`) used for model setup and training are made available (Supplementary Data 10).

## Data Availability

Scientists can obtain access to individual-level data from the UK Biobank by applying to UK Biobank (https://www.ukbiobank.ac.uk/enable-your-research).

## AUTHOR CONTRIBUTIONS

S.R. conceived the study and guided the analysis with D.W., S.B., O.W., O.L. and B.J.V. A.I.S. wrote the software and performed the analyses. K.R. assisted with literature review. A.I.S, B.J.V. and S.R. wrote the manuscript with contributions from all coauthors. All authors read and approved the final version of the manuscript.

## CODE AVAILABILITY

The code for EIR are available under the AGPL-v3 license and can be found at https://github.com/ arnor-sigurdsson/EIR and https://github.com/RasmussenLab/EIR.

## SUPPLEMENTARY INFORMATION

**Supplementary Figure 1.**
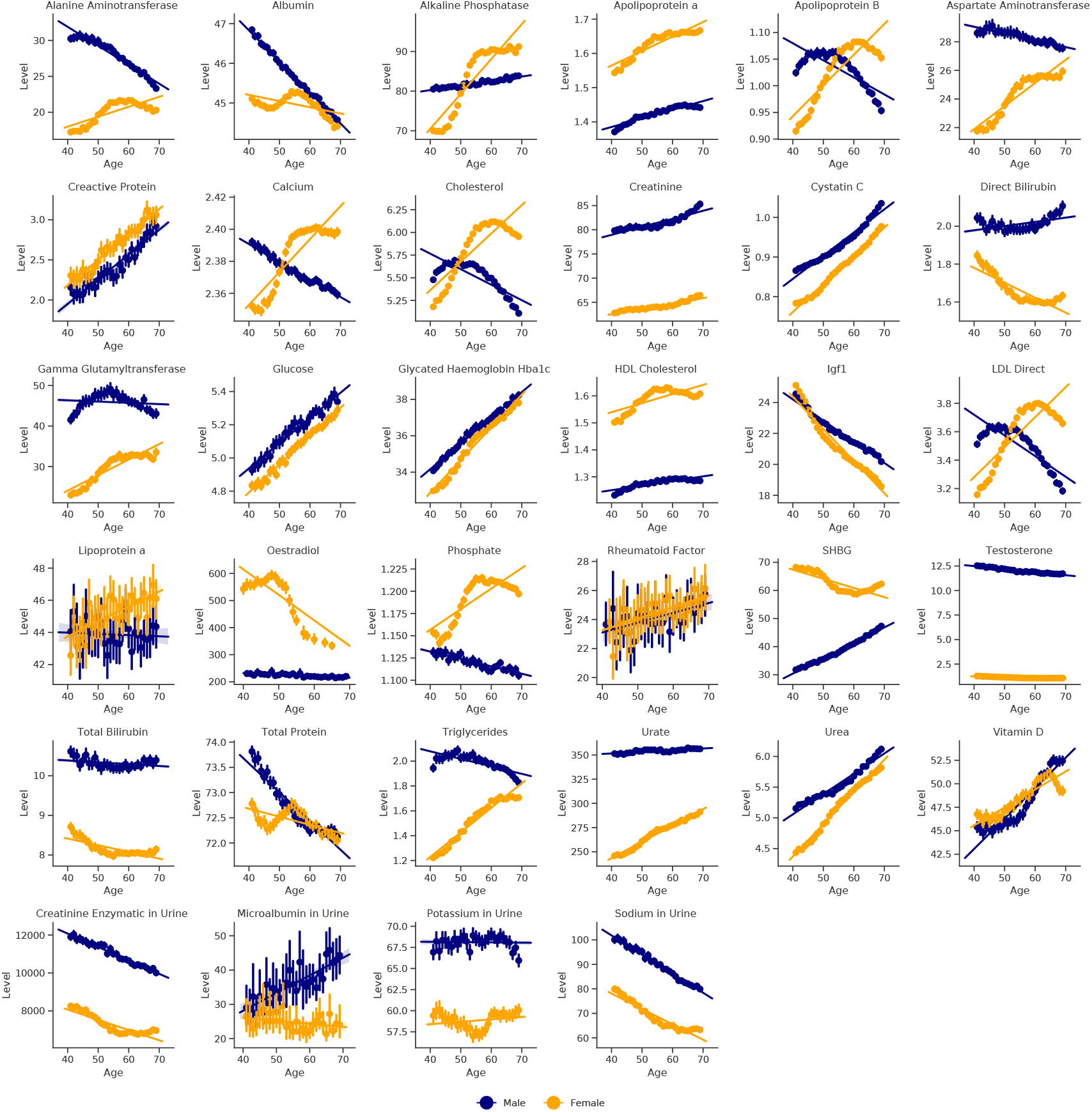
Effect of age and sex on all biomarker levels. The levels are plotted as a function of age, stratified by males (dark blue) and females (orange). Bars represent the 95% confidence interval after 1,000 bootstrap iterations. **Alanine Aminotransferase**: U/L, **Albumin**: g/L, **Alkaline Phosphatase**: U/L, **Apolipoprotein A**: g/L, **Apolipoprotein B**: g/L, **Aspartate Aminotransferase**: U/L, **Creactive Protein**: mg/L, **Calcium**: mmol/L, **Cholesterol**: mmol/L, **Creatinine**: umol/L, **Cystatin C**: mg/L, **Direct Bilirubin**: umol/L, **Gamma Glutamyltransferase**: U/L, **Glucose**: mmol/L, **Glycated Haemoglobin (HbA1c)**: mmol/mol, **HDL Cholesterol**: mmol/L, **IGF1**: nmol/L, **LDL Direct**: mmol/L, **Lipoprotein A**: nmol/L, **Oestradiol**: pmol/L, **Phosphate**: mmol/L, **Rheumatoid Factor**: IU/ml, **SHBG**: nmol/L, **Testosterone**: nmol/L, **Total Bilirubin**: umol/L, **Total Protein**: g/L, **Triglycerides**: mmol/L, **Urate**: umol/L, **Urea**: mmol/L, **Vitamin D**: nmol/L, **Creatinine Enzymatic in Urine**: umol/L, **Microalbumin in Urine**: mg/L, **Potassium in Urine**: mmol/L, **Sodium in Urine**: mmol/L.

**Supplementary Figure 2.**
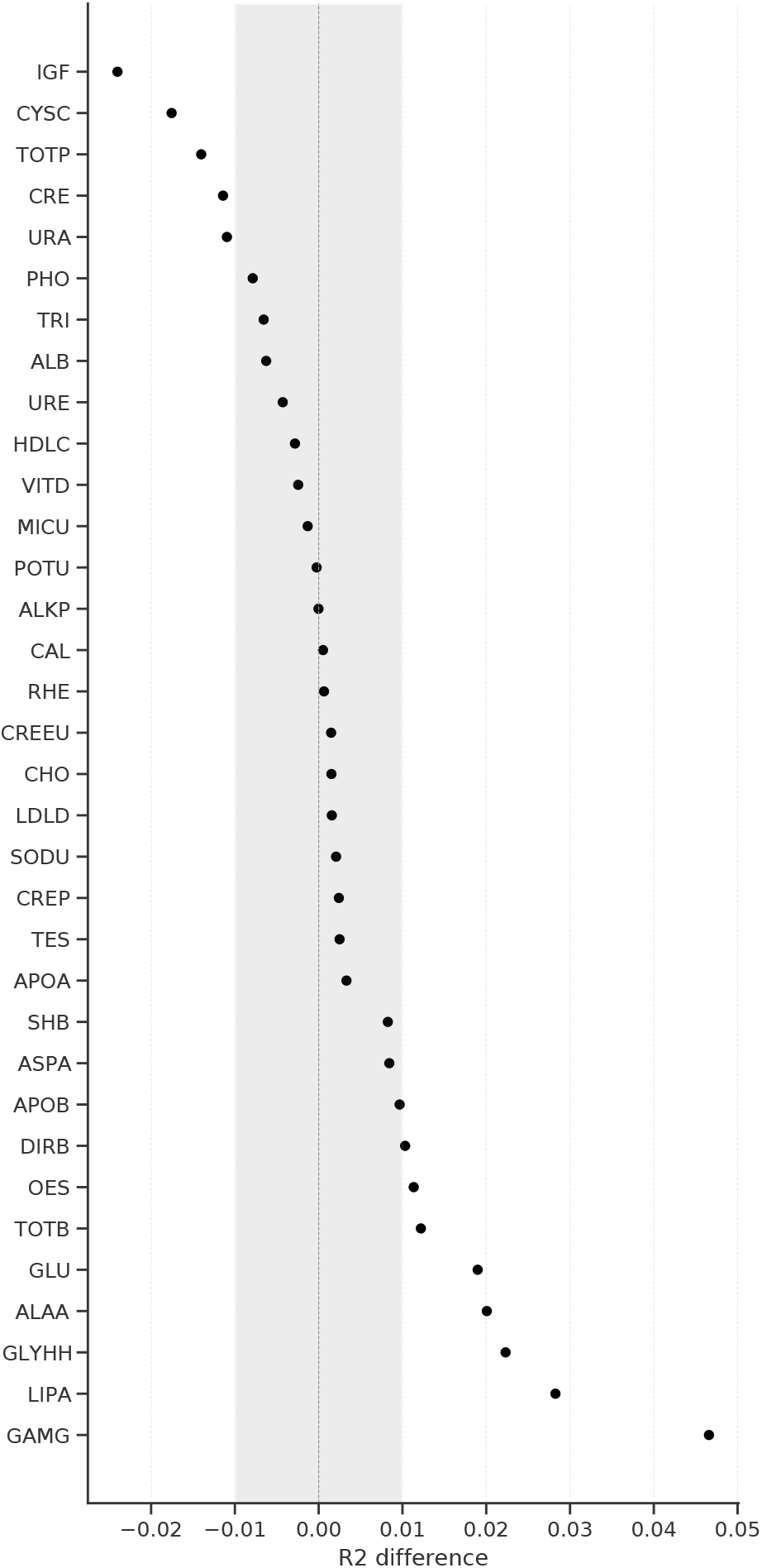
Comparison of SNPs used for deep-learning training. Difference in deep-learning genome-local-net (GLN) performance when using a set of 487,156 SNPs compared to pre-filtering SNPs based on genome-wide-association-studies (GWAS) performed on the training and validation set only, which resulted in 9,034 SNPs when using a p-value threshold of (5 *×* 10^−8^)*/*34. Negative numbers indicate that the full set performed better, while positive numbers indicate that the GWAS set performed better. Worse performance of the GWAS set indicates that the p-value threshold was likely too strict, relevant SNPs were filtered out. A better performance of the GWAS set indicates that the smaller number of features likely reduced overfitting when training the GLN models. The gray area is arbitrarily drawn at 0.0 ± 0.01 *R*^2^ for visual purposes. **TES**: Testosterone, **LIPA**: Lipoprotein A, **TOTB**: Total Bilirubin, **URA**: Urate, **DIRB**: Direct Bilirubin, **HDLC**: HDL Cholesterol, **CRE**: Creatinine, **SHB**: SHBG, **APOA**: Apolipoprotein A, **CYSC**: Cystatin C, **IGF**: IGF1, **ALKP**: Alkaline Phosphatase, **APOB**: Apolipoprotein B, **CHO**: Cholesterol, **TRI**: Triglycerides, **LDLD**: LDL Direct, **PHO**: Phosphate, **CREEU**: Creatinine Enzymatic in Urine, **URE**: Urea, **ALB**: Albumin, **OES**: Oestradiol, **ALAA**: Alanine Aminotransferase, **TOTP**: Total Protein, **GLYHH**: Glycated Haemoglobin Hba1c,**CAL**: Calcium **SODU**: Sodium in Urine, **VITD**: Vitamin D, **ASPA**: Aspartate Aminotransferase, **CREP**: Creactive Protein, **GAMG**: Gamma Glutamyltransferase, **POTU**: Potassium in Urine, **GLU**: Glucose, **MICU**: Microalbumin in Urine, **RHE**: Rheumatoid Factor.

**Supplementary Figure 3.**
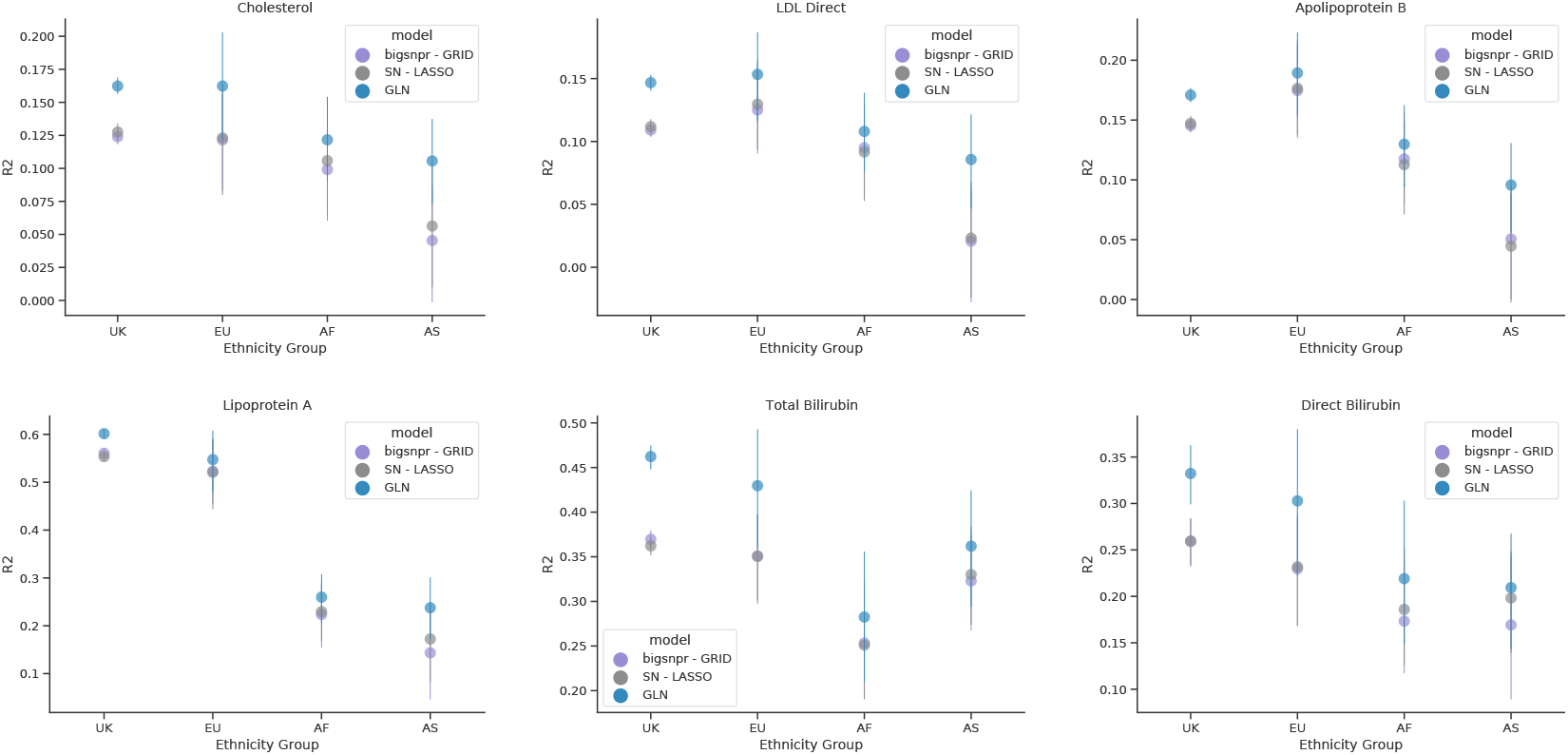
Performance in different populations. Performance in *R*^2^, stratified by self-reported continent of origin on the full 487,156 SNP set for the six traits where DL outperformed additive models. **UK**: United Kingdom, **EU**: Non-UK Europe, **AF**: Africa, **AS**: Asia. Bars represent the 95% CI from 1,000 bootstrap replicates on the held-out test set.

**Supplementary Figure 4.**
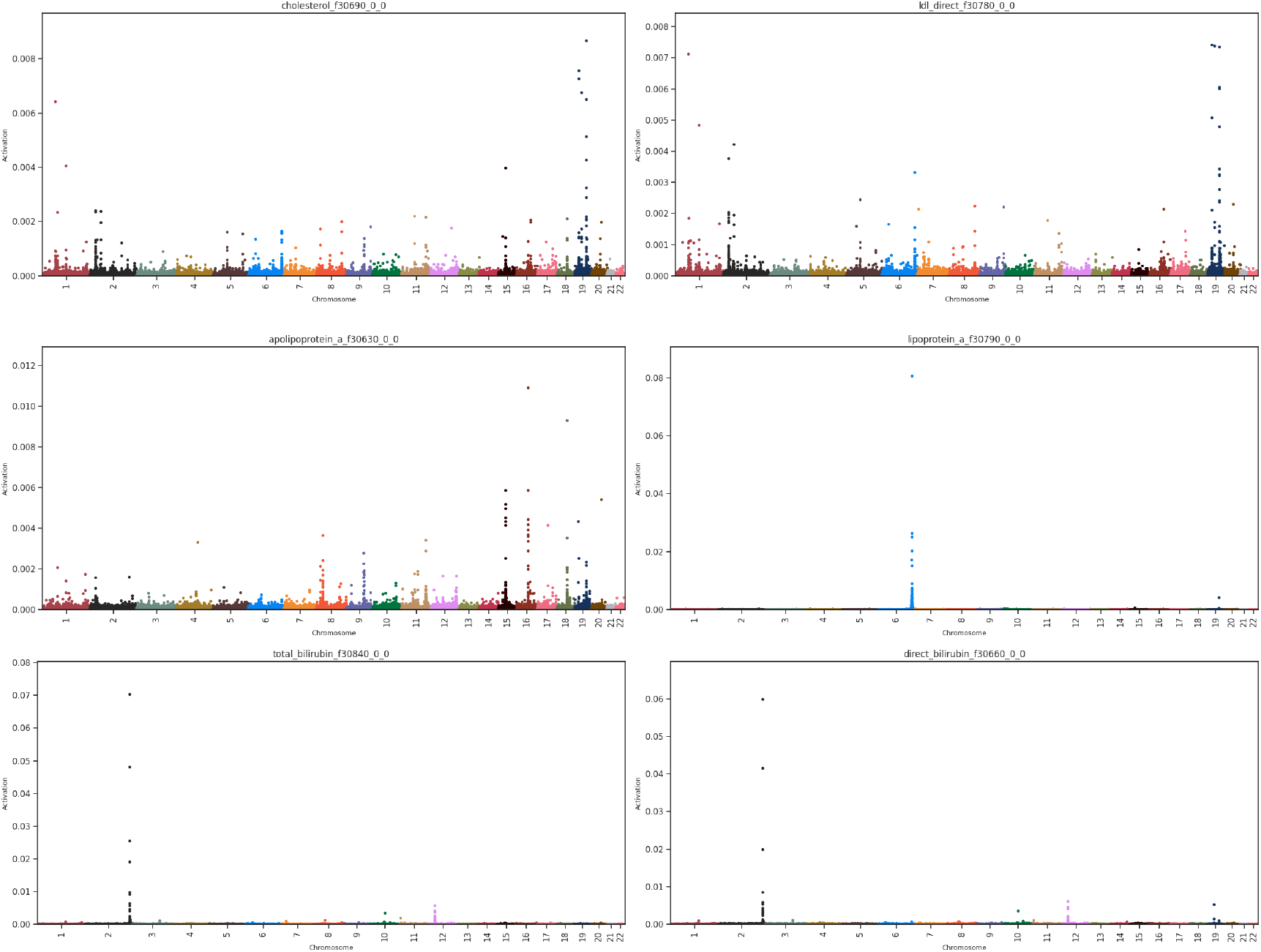
Deep learning genotype effects. SNP activation distribution for the six traits where DL performed favorably compared to the additive models. The y-axis represents average SHAP activation for the DL GLN model per SNP. Each point represents a variant, and the points are colored according to chromosomes.

**Supplementary Figure 5.**
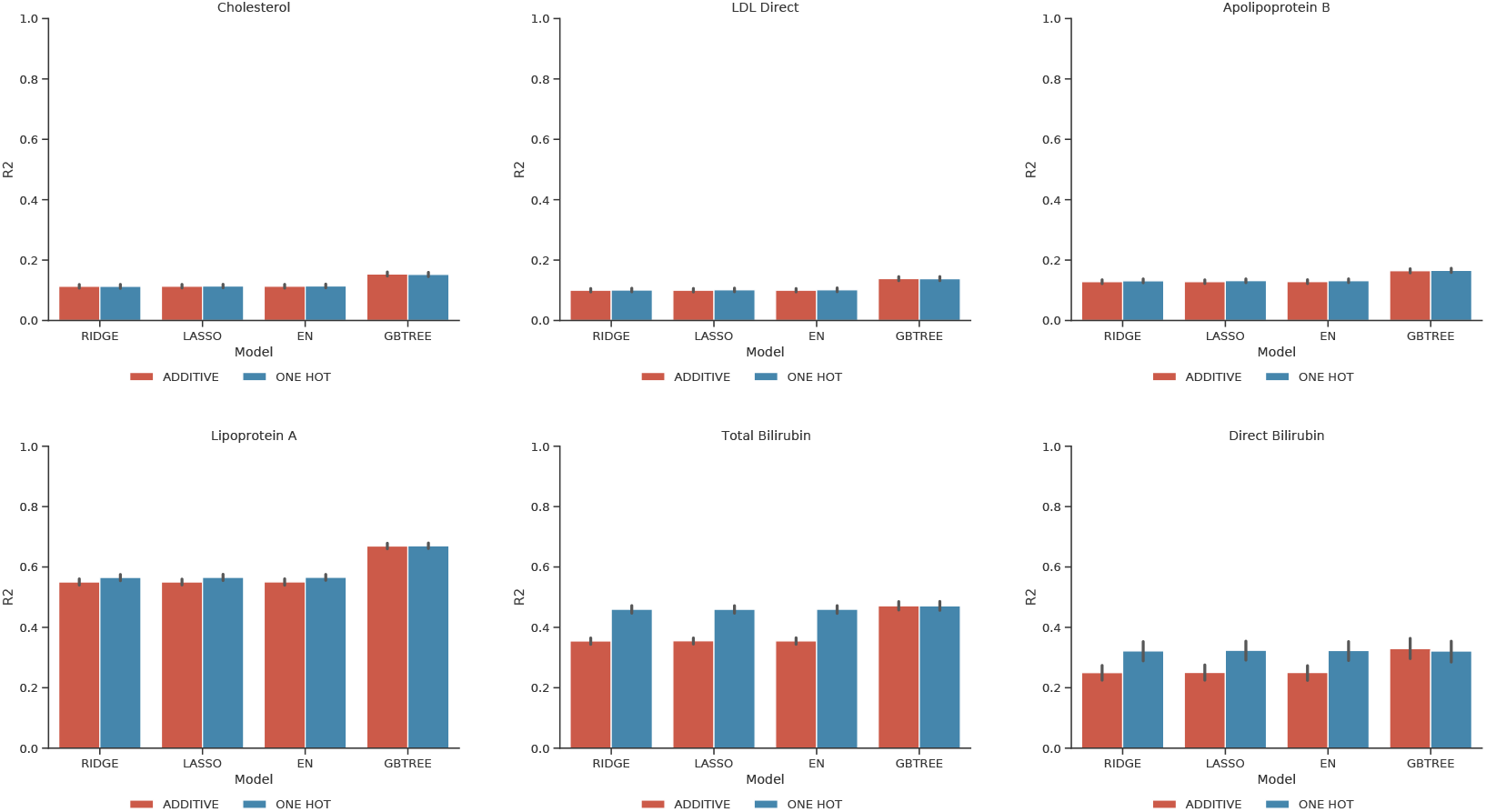
Performance comparison of linear and non-linear models, trained on samples of self-reported UK origin, evaluated on samples with self-reported UK origin. Models were trained on both additive and one-hot (each allele modelled separately) encodings of the genotype data. For each trait, a subset of SNPs most highly activated by the genome-local-net (GLN) model on the validation set were used for training and evaluation. All models were trained on data encoded with an additive prior (dark blue) and in a one-hot encoding (red), where each allele can be modelled separately. Bars represent the 95% CI from 1,000 bootstrap replicates on the held-out test set. **RIDGE**: Ridge Regression, **LASSO**: Lasso Regression, **EN**: Elastic-Net regression, **GBTREE**: Gradient Boosted Decision Trees Regression.

**Supplementary Figure 6.**
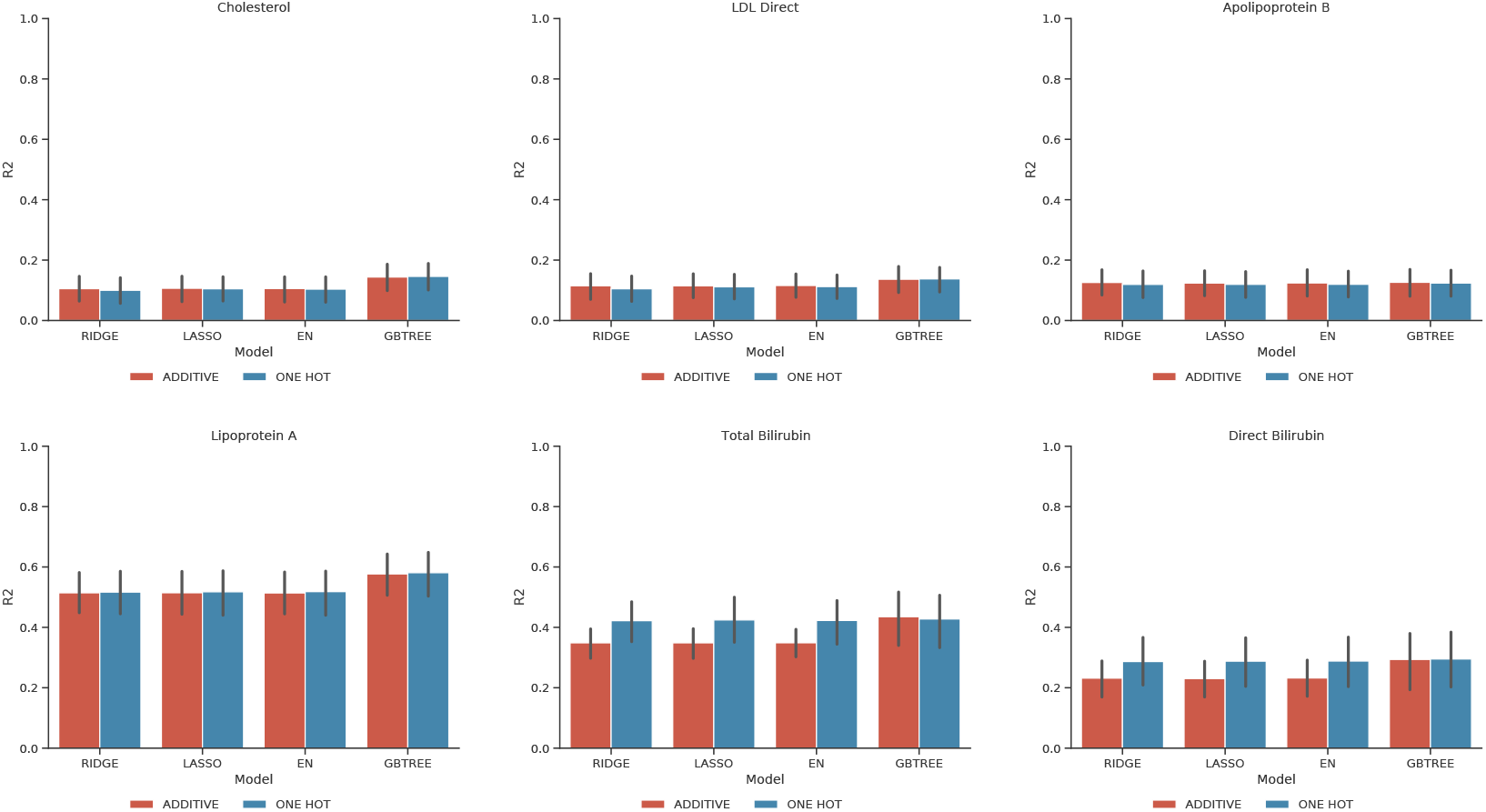
Performance comparison of linear and non-linear models, trained on samples of self-reported UK origin, evaluated on samples with self-reported non-UK European origin. Models were trained on both additive and one-hot (each allele modelled separately) encodings of the genotype data. For each trait, a subset of SNPs most highly activated by the genome-local-net (GLN) model on the validation set were used for training and evaluation. All models were trained on data encoded with an additive prior (dark blue) and in a one-hot encoding (red), where each allele can be modelled separately. Bars represent the 95% CI from 1,000 bootstrap replicates on the held-out test set. **RIDGE**: Ridge Regression, **LASSO**: Lasso Regression, **EN**: Elastic-Net regression, **GBTREE**: Gradient Boosted Decision Trees Regression.

**Supplementary Figure 7.**
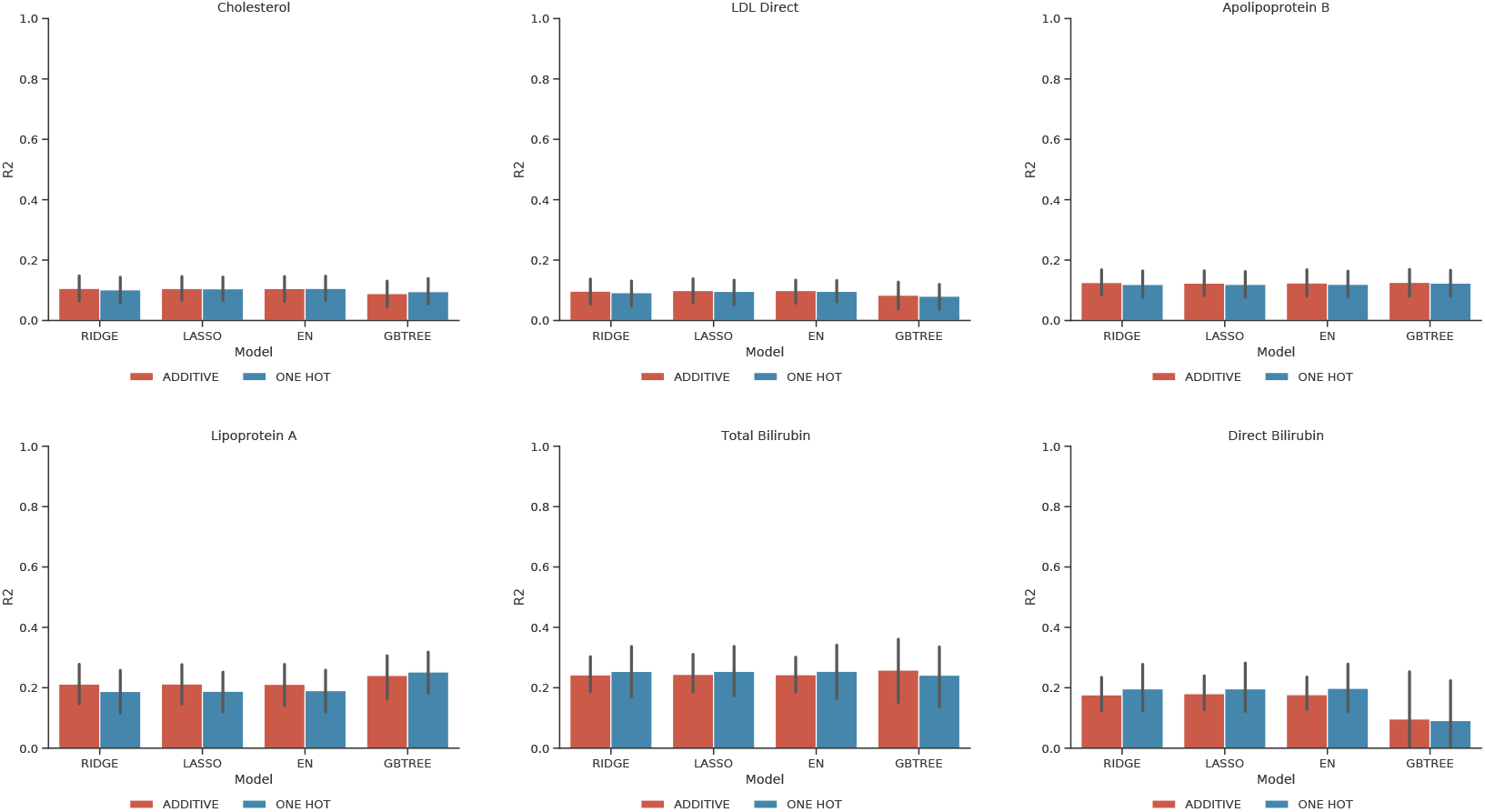
Performance comparison of linear and non-linear models, trained on samples of self-reported UK origin, evaluated on samples with self-reported African origin. Models were trained on both additive and one-hot (each allele modelled separately) encodings of the genotype data. For each trait, a subset of SNPs most highly activated by the genome-local-net (GLN) model on the validation set were used for training and evaluation. All models were trained on data encoded with an additive prior (dark blue) and in a one-hot encoding (red), where each allele can be modelled separately. Bars represent the 95% CI from 1,000 bootstrap replicates on the held-out test set. **RIDGE**: Ridge Regression, **LASSO**: Lasso Regression, **EN**: Elastic-Net regression, **GBTREE**: Gradient Boosted Decision Trees Regression.

**Supplementary Figure 8.**
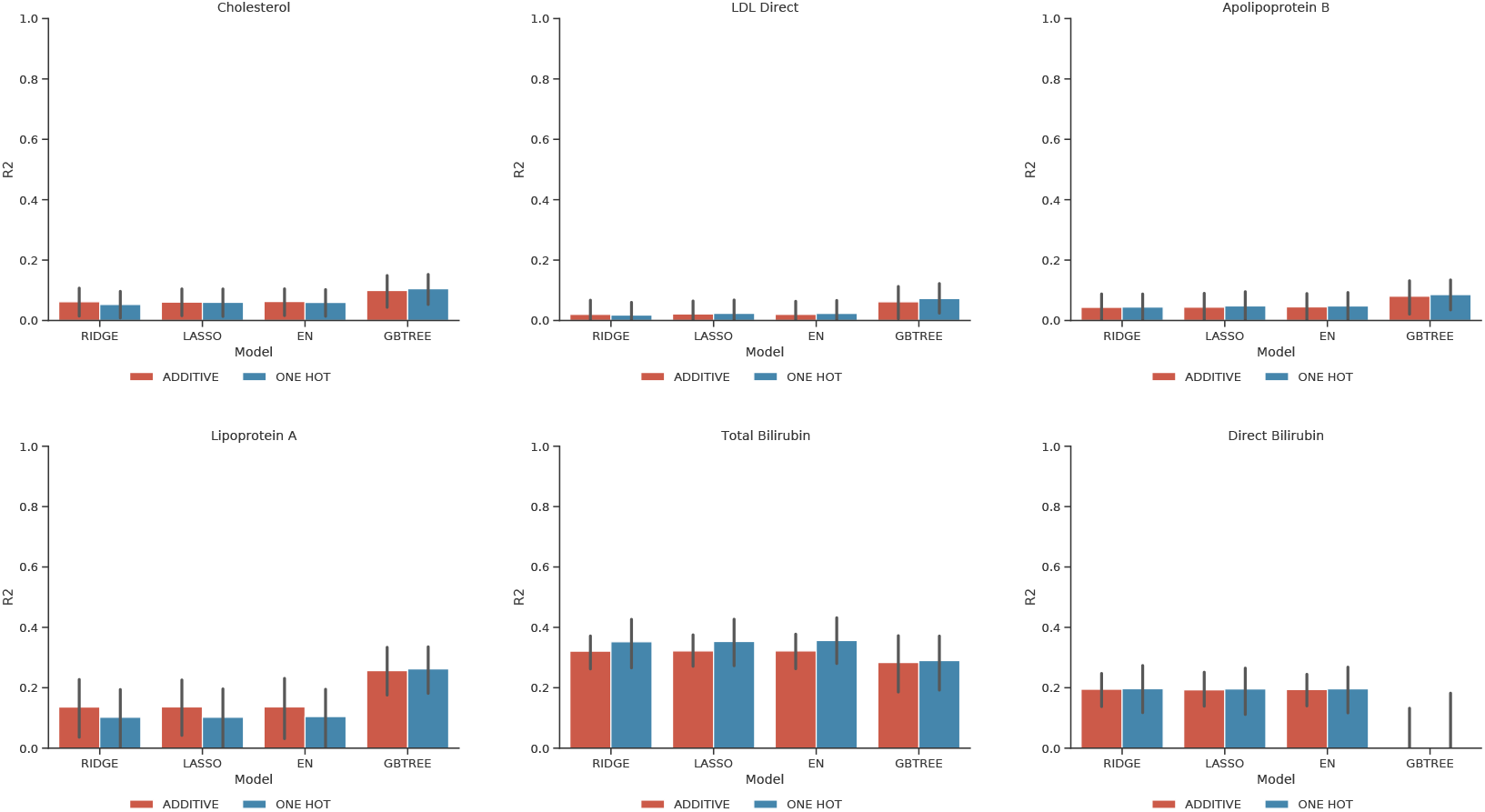
Performance comparison of linear and non-linear models, trained on samples of self-reported UK origin, evaluated on samples with self-reported Asian origin. Models were trained on both additive and one-hot (each allele modelled separately) encodings of the genotype data. For each trait, a subset of SNPs most highly activated by the genome-local-net (GLN) model on the validation set were used for training and evaluation. All models were trained on data encoded with an additive prior (dark blue) and in a one-hot encoding (red), where each allele can be modelled separately. Bars represent the 95% CI from 1,000 bootstrap replicates on the held-out test set. **RIDGE**: Ridge Regression, **LASSO**: Lasso Regression, **EN**: Elastic-Net regression, **GBTREE**: Gradient Boosted Decision Trees Regression.

**Supplementary Figure 9.**
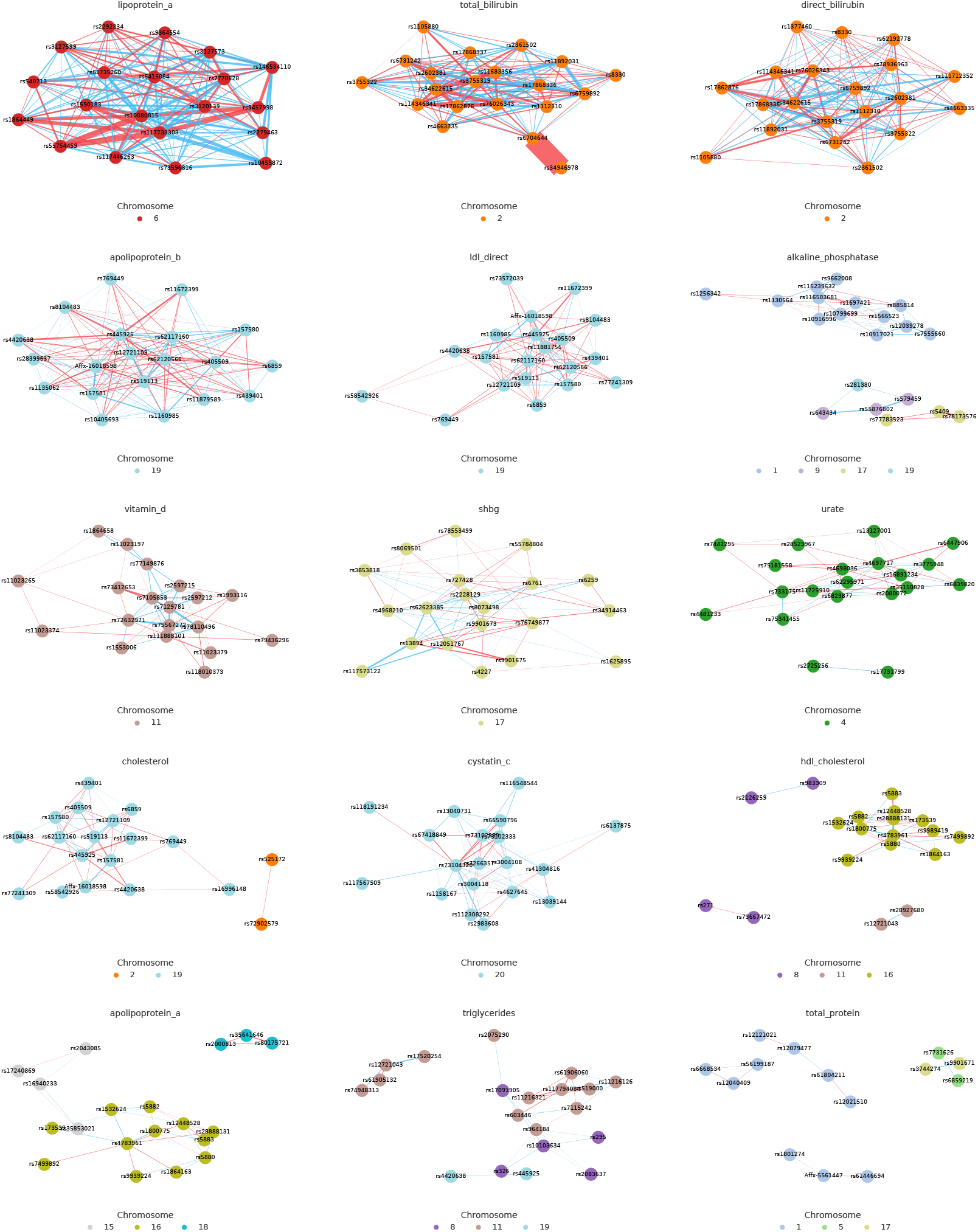
Traits with highest identified interaction effects. Shown are the 15 traits with the highest total interaction effects. Computing the interactions between SNPs with an ordinary-least-squares (OLS) model finds numerous interaction effects, which vary in strength and number depending on trait. Shown are the top 20 SNPs with the highest total interaction effects for each trait. Each node represents a SNP, and the edges reflect interaction effects identified by the OLS model affecting a biomarker level. The edges a colored according to whether the interaction effect increases (red) or decreases (blue) the biomarker level, and the width of the edges reflect the relative strength of the interaction between SNPs. Nodes are colored by chromosome. Note that since only the top 20 SNPs are shown, many effects are missing from the plot.

**Supplementary Table 1.** Results when fitting an ordinary least squares (OLS) model on chosen SNPs for specific biomarkers in the blood. The results indicate that strong non-additive effects modulate the amount of certain biomarkers in the blood.

| Allele | $\beta$ | STDERR | $t$ | $p$ | 2.5% CI | 97.5% CI | Trait |
| --- | --- | --- | --- | --- | --- | --- | --- |
| rs10455872 AA | -0.2533 | 0.002 | -158.021 | 0.0 | -0.256 | -0.25 | Lipoprotein A |
| rs10455872 AG | 1.5839 | 0.004 | 391.968 | 0.0 | 1.576 | 1.592 | Lipoprotein A |
| rs10455872 GG | 1.9708 | 0.035 | 55.866 | 0.0 | 1.902 | 2.04 | Lipoprotein A |
| rs887829 CC | -0.3613 | 0.002 | -180.271 | 0.0 | -0.365 | -0.357 | Total Bilirubin |
| rs887829 CT | -0.0181 | 0.002 | -8.728 | 0.0 | -0.022 | -0.014 | Total Bilirubin |
| rs887829 TT | 1.7505 | 0.004 | 404.585 | 0.0 | 1.742 | 1.759 | Total Bilirubin |

## REFERENCES

1. Qiu, X. et al. The diagnostic value of serum creatinine and cystatin c in evaluating glomerular filtration rate in patients with chronic kidney disease: a systematic literature review and meta-analysis. Oncotarget 8, 72985–72999 (2017). URL https://www.ncbi.nlm.nih.gov/pmc/articles/PMC5641185/.

2. Spinella, R., Sawhney, R. & Jalan, R. Albumin in chronic liver disease: structure, functions and therapeutic implications. Hepatology International 10, 124–132 (2016).

3. Carson, J. A. S. et al. Dietary Cholesterol and Cardiovascular Risk: A Science Advisory From the American Heart Association. Circulation 141, e39–e53 (2020). URL https://www.ahajournals.org/doi/full/10.1161/CIR.0000000000000743.

4. Reyes-Soffer, G. et al. Lipoprotein(a): A Genetically Determined, Causal, and Prevalent Risk Factor for Atherosclerotic Cardiovascular Disease: A Scientific Statement From the American Heart Association. Arteriosclerosis, Thrombosis, and Vascular Biology 42, e48–e60 (2022). URL https://www.ahajournals.org/doi/10.1161/ATV.0000000000000147.

5. Caplice, N. M. et al. Lipoprotein (a) binds and inactivates tissue factor pathway inhibitor: a novel link between lipoproteins and thrombosis. Blood 98, 2980–2987 (2001).

6. Smolders, B., Lemmens, R. & Thijs, V. Lipoprotein (a) and Stroke: A Meta-Analysis of Observational Studies. Stroke 38, 1959–1966 (2007). URL https://www.ahajournals.org/doi/10.1161/STROKEAHA.106.480657.

7. Stone, N. J. et al. 2013 ACC/AHA Guideline on the Treatment of Blood Cholesterol to Reduce Atherosclerotic Cardiovascular Risk in Adults. Journal of the American College of Cardiology 63, 2889–2934 (2014). URL https://linkinghub.elsevier.com/retrieve/pii/S0735109713060282.

8. Basu, N. N. et al. Risk of contralateral breast cancer in BRCA1 and BRCA2 mutation carriers: a 30-year semi-prospective analysis. Familial Cancer 14, 531–538 (2015). URL http://link.springer.com/10.1007/s10689-015-9825-9.

9. Wasung, M. E., Chawla, L. S. & Madero, M. Biomarkers of renal function, which and when? Clinica Chimica Acta 438, 350–357 (2015). URL https://linkinghub.elsevier.com/retrieve/pii/S000989811400391X.

10. Davis, K. D. et al. Discovery and validation of biomarkers to aid the development of safe and effective pain therapeutics: challenges and opportunities. Nature Reviews Neurology 16, 381–400 (2020). URL https://www.nature.com/articles/s41582-020-0362-2.

11. Chartier-Hariln, M.-C. et al. Apolipoprotein E, e4 allele as a major risk factor for sporadic early and late-onset forms of Alzheimer’s disease: analysis of the 19q13.2 chromosomal region. Human Molecular Genetics 3, 569–574 (1994). URL https://academic.oup.com/hmg/article-lookup/doi/10.1093/hmg/3.4.569.

12. Ledermann, J. et al. Olaparib Maintenance Therapy in Platinum-Sensitive Relapsed Ovarian Cancer. New England Journal of Medicine 366, 1382–1392 (2012). URL http://www.nejm.org/doi/abs/10.1056/NEJMoa1105535.

13. Wallace, C. et al. Genome-wide association study identifies genes for biomarkers of cardiovascular disease: serum urate and dyslipidemia. American Journal of Human Genetics 82, 139–149 (2008).

14. Johnson, A. D. et al. Genome-wide association meta-analysis for total serum bilirubin levels. Human Molecular Genetics 18, 2700–2710 (2009). URL https://academic.oup.com/hmg/article-lookup/doi/10.1093/hmg/ddp202.

15. Schmidt, K., Noureen, A., Kronenberg, F. & Utermann, G. Structure, function, and genetics of lipoprotein (a). Journal of Lipid Research 57, 1339–1359 (2016). URL https://www.jlr.org/article/S0022-2275(20)35208-1/abstract.

16. Sinnott-Armstrong, N. et al. Genetics of 35 blood and urine biomarkers in the UK Biobank. Nature Genetics 53, 185–194 (2021). URL https://www.nature.com/articles/s41588-020-00757-z.

17. Polygenic Risk Score Task Force of the International Common Disease Alliance et al. Responsible use of polygenic risk scores in the clinic: potential benefits, risks and gaps. Nature Medicine 27, 1876–1884 (2021). URL https://www.nature.com/articles/s41591-021-01549-6.

18. Lewis, A. C. F. & Green, R. C. Polygenic risk scores in the clinic: new perspectives needed on familiar ethical issues. Genome Medicine 13, 14 (2021). URL https://genomemedicine.biomedcentral.com/articles/10.1186/s13073-021-00829-7.

19. Torkamani, A., Wineinger, N. E. & Topol, E. J. The personal and clinical utility of polygenic risk scores. Nature Reviews Genetics 19, 581–590 (2018). URL https://www.nature.com/articles/s41576-018-0018-x.

20. Lambert, S. A., Abraham, G. & Inouye, M. Towards clinical utility of polygenic risk scores. Human Molecular Genetics 28, R133–R142 (2019). URL https://doi.org/10.1093/hmg/ddz187.

21. Lewis, C. M. & Vassos, E. Polygenic risk scores: from research tools to clinical instruments. Genome Medicine 12, 44 (2020). URL https://doi.org/10.1186/s13073-020-00742-5.

22. Kullo, I. J. et al. Polygenic scores in biomedical research. Nature Reviews Genetics 23, 524–532 (2022). URL https://www.nature.com/articles/s41576-022-00470-z.

23. Cho, S. M. J. et al. Measured Blood Pressure, Genetically Predicted Blood Pressure, and Cardiovascular Disease Risk in the UK Biobank. JAMA Cardiology (2022). URL https://doi.org/10.1001/jamacardio.2022.3191.

24. Loh, P.-R. et al. Efficient Bayesian mixed-model analysis increases association power in large cohorts. Nature Genetics 47, 284–290 (2015). URL http://www.nature.com/articles/ng.3190.

25. Moser, G. et al. Simultaneous Discovery, Estimation and Prediction Analysis of Complex Traits Using a Bayesian Mixture Model. PLOS Genetics 11, e1004969 (2015). URL https://journals.plos.org/plosgenetics/article?id=10.1371/journal.pgen.1004969.

26. Privé, F., Aschard, H., Ziyatdinov, A. & Blum, M. G. B. Efficient analysis of large-scale genome-wide data with two R packages: bigstatsr and bigsnpr. Bioinformatics 34, 2781–2787 (2018). URL https://academic.oup.com/bioinformatics/article/34/16/2781/4956666.

27. Qian, J. et al. A fast and scalable framework for large-scale and ultrahigh-dimensional sparse regression with application to the UK Biobank. PLOS Genetics 16, e1009141 (2020). URL https://journals.plos.org/plosgenetics/article?id=10.1371/journal.pgen.1009141.

28. Li, R. et al. Fast numerical optimization for genome sequencing data in population biobanks. Bioinformatics 37, 4148–4155 (2021). URL https://doi.org/10.1093/bioinformatics/btab452. https://academic.oup.com/bioinformatics/article-pdf/37/22/4148/45966467/btab452.pdf.

29. Vilhjálmsson, B. et al. Modeling Linkage Disequilibrium Increases Accuracy of Polygenic Risk Scores. The American Journal of Human Genetics 97, 576–592 (2015). URL https://linkinghub.elsevier.com/retrieve/pii/S0002929715003651.

30. Mak, T. S. H., Porsch, R. M., Choi, S. W., Zhou, X. & Sham, P. C. Polygenic scores via penalized regression on summary statistics. Genetic Epidemiology 41, 469–480 (2017). URL https://onlinelibrary.wiley.com/doi/abs/10.1002/gepi.22050.

31. Lloyd-Jones, L. R. et al. Improved polygenic prediction by Bayesian multiple regression on summary statistics. Nature Communications 10, 5086 (2019). URL http://www.nature.com/articles/s41467-019-12653-0.

32. Ge, T., Chen, C.-Y., Ni, Y., Feng, Y.-C. A. & Smoller, J. W. Polygenic prediction via Bayesian regression and continuous shrinkage priors. Nature Communications 10, 1776 (2019). URL http://www.nature.com/articles/s41467-019-09718-5.

33. Privé, F., Arbel, J. & Vilhjálmsson, B. J. LDpred2: better, faster, stronger. Bioinformatics 36, 5424–5431 (2020). URL https://doi.org/10.1093/bioinformatics/btaa1029.

34. Privé, F., Vilhjálmsson, B. J. & Mak, T. S. H. lassosum2: an updated version complementing LDpred2. bioRxiv 2021.03.29.437510 (2021). URL https://www.biorxiv.org/content/10.1101/2021.03.29.437510v1.

35. Bellot, P., de los Campos, G. & Pérez-Enciso, M. Can Deep Learning Improve Genomic Prediction of Complex Human Traits? Genetics 210, 809–819 (2018). URL https://doi.org/10.1534/genetics.118.301298.

36. Xu, Y. et al. Learning polygenic scores for human blood cell traits. bioRxiv 2020.02.17.952788 (2020). URL https://www.biorxiv.org/content/10.1101/2020.02.17.952788v1.

37. Sigurdsson, A. I. et al. Deep integrative models for large-scale human genomics. bioRxiv (2021). URL https://www.biorxiv.org/content/early/2021/09/03/2021.06.11.447883. https://www.biorxiv.org/content/early/2021/09/03/2021.06.11.447883.full.pdf.

38. Elgart, M. et al. Non-linear machine learning models incorporating SNPs and PRS improve polygenic prediction in diverse human populations. Communications Biology 5, 1–12 (2022). URL https://www.nature.com/articles/s42003-022-03812-z.

39. Albiñana, C. et al. Multi-PGS enhances polygenic prediction: weighting 937 polygenic scores (2022). URL https://www.medrxiv.org/content/10.1101/2022.09.14.22279940v1.

40. Bycroft, C. et al. The UK Biobank resource with deep phenotyping and genomic data. Nature 562, 203–209 (2018). URL http://www.nature.com/articles/s41586-018-0579-z.

41. Lundberg, S. M. et al. From local explanations to global understanding with explainable AI for trees. Nature Machine Intelligence 2, 56–67 (2020). URL http://www.nature.com/articles/s42256-019-0138-9.

42. Lundberg, S. M. & Lee, S.-I. A Unified Approach to Interpreting Model Predictions. Advances in Neural Information Processing Systems 30 (2017). URL https://papers.nips.cc/paper/2017/hash/8a20a8621978632d76c43dfd28b67767-Abstract.html.

43. Cardoso-Saldaña, G. et al. The rs10455872-g allele of the lpa gene is associated with high lipoprotein levels and increased aortic valve calcium in a mexican adult population. Genetics and Molecular Biology 42, 519–525 (2019).

44. Page, M. M. et al. Coronary artery disease and the risk-associated lpa variants, rs3798220 and rs10455872, in patients with suspected familial hypercholesterolaemia. Clinica Chimica Acta 510, 211–215 (2020).

45. Horsfall, L. J., Burgess, S., Hall, I. & Nazareth, I. Genetically raised serum bilirubin levels and lung cancer: a cohort study and mendelian randomisation using uk biobank. Thorax 75, 955–964 (2020).

46. Zeng, L. et al. Cis-epistasis at the LPA locus and risk of cardiovascular diseases. Cardiovascular Research 118, 1088–1102 (2022).

47. Novakovsky, G., Dexter, N., Libbrecht, M. W., Wasserman, W. W. & Mostafavi, S. Obtaining genetics insights from deep learning via explainable artificial intelligence. Nature Reviews Genetics 1–13 (2022). URL https://www.nature.com/articles/s41576-022-00532-2.

48. Acosta, J. N., Falcone, G. J., Rajpurkar, P. & Topol, E. J. Multimodal biomedical AI. Nature Medicine 28, 1773–1784 (2022). URL https://www.nature.com/articles/s41591-022-01981-2.

49. Purcell, S. et al. PLINK: A Tool Set for Whole-Genome Association and Population-Based Linkage Analyses. American Journal of Human Genetics 81, 559–575 (2007). URL https://www.ncbi.nlm.nih.gov/pmc/articles/PMC1950838/.

50. Paszke, A. et al. PyTorch: An Imperative Style, High-Performance Deep Learning Library. Advances in Neural Information Processing Systems 32 (2019). URL https://papers.nips.cc/paper/2019/hash/bdbca288fee7f92f2bfa9f7012727740-Abstract.html.

51. Kingma, D. P. & Ba, J. Adam: A Method for Stochastic Optimization. 1412.6980 [cs] (2017). URL http://arxiv.org/abs/1412.6980. ArXiv: 1412.6980.

52. Hendrycks, D. & Gimpel, K. Gaussian Error Linear Units (GELUs). 1606.08415 [cs] (2020). URL http://arxiv.org/abs/1606.08415. ArXiv: 1606.08415.

53. Elfwing, S., Uchibe, E. & Doya, K. Sigmoid-weighted linear units for neural network function approximation in reinforcement learning. Neural Networks 107, 3–11 (2018). URL https://www.sciencedirect.com/science/article/pii/S0893608017302976. Special issue on deep reinforcement learning.

54. Ramachandran, P., Zoph, B. & Le, Q. V. Searching for Activation Functions. arXiv:1710.05941 [cs](2017). URL http://arxiv.org/abs/1710.05941. ArXiv: 1710.05941.

55. Zhang, H., Cisse, M., Dauphin, Y. N. & Lopez-Paz, D. mixup: Beyond Empirical Risk Minimization. arXiv:1710.09412 [cs, stat] (2018). URL http://arxiv.org/abs/1710.09412. ArXiv: 1710.09412 version: 2.

56. Yun, S. et al. CutMix: Regularization Strategy to Train Strong Classifiers With Localizable Features. In 2019 IEEE/CVF International Conference on Computer Vision (ICCV), 6022–6031 (2019). ISSN: 2380-7504.

57. Chen, T. & Guestrin, C. XGBoost: A scalable tree boosting system. In Proceedings of the 22nd ACM SIGKDD International Conference on Knowledge Discovery and Data Mining, KDD ‘16, 785–794 (ACM, New York, NY, USA, 2016). URL http://doi.acm.org/10.1145/2939672.2939785.

58. He, K., Zhang, X., Ren, S. & Sun, J. Deep Residual Learning for Image Recognition. In 2016 IEEE Conference on Computer Vision and Pattern Recognition (CVPR), 770–778 (2016). ISSN: 1063-6919.

59. Ngiam, J. et al. Tiled convolutional neural networks. In Lafferty, J., Williams, C., Shawe-Taylor, J., Zemel, R. & Culotta, A. (eds.) Advances in Neural Information Processing Systems, vol. 23 (Curran Associates, Inc., 2010). URL https://proceedings.neurips.cc/paper/2010/file/01f78be6f7cad02658508fe4616098a9-Paper.pdf.

60. Taigman, Y., Yang, M., Ranzato, M. & Wolf, L. DeepFace: Closing the Gap to Human-Level Performance in Face Verification. In 2014 IEEE Conference on Computer Vision and Pattern Recognition, 1701–1708 (2014). ISSN: 1063-6919.

61. Chen, Y.-h. et al. Locally-connected and convolutional neural networks for small footprint speaker recognition. In Sixteenth Annual Conference of the International Speech Communication Association (2015).

62. Srivastava, N., Hinton, G., Krizhevsky, A., Sutskever, I. & Salakhutdinov, R. Dropout: a simple way to prevent neural networks from overfitting. The Journal of Machine Learning Research 15, 1929–1958 (2014).

63. Huang, G., Sun, Y., Liu, Z., Sedra, D. & Weinberger, K. Q. Deep Networks with Stochastic Depth. In Leibe, B., Matas, J., Sebe, N. & Welling, M. (eds.) Computer Vision – ECCV 2016, Lecture Notes in Computer Science, 646–661 (Springer International Publishing, Cham, 2016).

